# A qualitative investigation of crisis cafés in England: their role, implementation, and accessibility

**DOI:** 10.1101/2024.05.09.24306304

**Authors:** Heather Staples, Gianna Cadorna, Patrick Nyikavaranda, Lucy Maconick, Brynmor Lloyd-Evans, Sonia Johnson

## Abstract

**Background:** Crisis cafés (also known as crisis sanctuaries or havens) are community-based services which support people in mental health crises, aiming to provide an informal, non-clinical and accessible setting. This model is increasingly popular in the UK; however, we are aware of no peer-reviewed literature focused on this model. We aimed to use qualitative methods to investigate managers’ views of the aims of crisis cafés, how they operate in practice and the factors that affect access to these services and implementation of the intended model.

**Methods:** Semi-structured interviews were conducted with 12 managers of crisis cafés across England. Data were analysed using a thematic approach.

**Results:** We identified five main perceived aims for crisis cafés: providing an alternative to A&E; improving access to crisis care; providing people in acute distress with someone to talk to in a safe and comfortable space; triaging effectively; and improving crisis planning and people’s coping skills. Factors seen as influencing the effectiveness of crisis cafés included accessibility, being able to deliver person-centred care, relationships with other services, and staffing. These factors could both help and hinder access to care and the implementation of the intended model. There were a number of trade-offs that services had to consider when designing and running a crisis café: 1. Balancing an open-door policy with managing demand for the service through referral routes, 2. Balancing risk management procedures with the remit of offering a non-clinical environment and 3. Increasing awareness of the service in the community whilst avoiding stigmatising perceptions of it.

**Conclusions:** Findings illustrate the aims of the crisis café model of care and factors which are influential in its implementation in current practice. Future research is needed to evaluate the efficacy of these services in relation to their aims. Crisis café service users’ views, and views of stakeholders from the wider crisis care system should also be ascertained.

## Background

A mental health crisis can occur when a person experiences severe distress or a breakdown in functioning that exceeds their capacity to cope, which can result in behaviour that poses significant risk to themselves or others [1, 2]. For many people experiencing mental health crises, presenting to Emergency Departments can seem to be the only option [3], especially when urgent help is needed outside regular working hours when other avenues of support are unavailable. However, the environment in these services is far from ideal for people in mental health crisis. Emergency Departments tend to be loud, hectic and expose many service users to sights and sounds which may increase their distress [4, 5]. Crisis resolution teams (CRTs), which are the main source of community crisis care, do not always provide a full in-person service 24 hours a day and are often not open to self-referrals [6]. Additionally, CRTs were established as an alternative to inpatient admission, so focus on people whose difficulties are severe enough for them to be at risk of admission [7]. This leaves a gap in service provision for people who are severely distressed, but do not quite meet the threshold for hospital admission and may not find the environments in an Emergency Department or in hospital helpful. For some people in severe distress, an emergency response that is less clinically focused may also be helpful [8].

Research has been conducted with service users to ascertain their views on crisis care. A very high priority among service users is to be treated in a warm, caring and respectful way [9]. Service users often describe negative experiences of presenting to Emergency Departments in mental health crisis, with variability in the availability of staff with specialist training in mental health, stigmatising attitudes towards mental health patients, and an uncomfortable environment to wait in [9, 10].

Particularly, difficulties getting access to effective help and negative attitudes from both general Emergency Department staff and those specialising in mental health are often reported for people whose presentations involve self-harm or a “personality disorder” diagnosis [11]. Many service users feel that an Emergency Department is not the right environment to receive mental health support when in crisis, instead looking to different services in the community [9, 10, 12]. As hospital admissions are costly, often unpopular and do not result in clear improvements in outcomes, a focus on developing alternatives to hospital and triaging people to them is mirrored in mental health policy and service development in many countries [5].

In England, the principal type of crisis service supported by policy over the past 20 years has been the crisis resolution and home treatment team, providing assessment and intensive treatment mainly in patients’ homes [13]. Even though many service users value the option of treatment at home, it is clear that these teams do not meet a full range of service user needs in crisis: they are intended to be narrowly focused on crises of a severity close to the threshold for hospital admission, and service users, especially those with a “personality disorder” diagnosis, often report that their needs for a readily accessible service in which they are able to form good therapeutic relationships with staff who have time to talk are not met [11, 14]. Crisis care improvement is currently a key policy target in England, with an aim that people should have access to good quality mental health crisis care ‘around the clock, 365 days a year’ [15, 16]. Growing awareness that one model is not likely to meet the full range of needs for alternatives to A & E attendance and to psychiatric hospital has driven a policy commitment that by 2023/24, every catchment area in England should offer a range of alternative services to A&E and to psychiatric admission within in all local crisis care pathways [17].

One form of alternative crisis care provision that has gained popularity over the recent years is the crisis café, sometimes also referred to as a crisis sanctuary, safe haven or recovery café [5]. According to Dalton-Locke et al. [8], a crisis café is “a community-based service which provides support for people in mental health crisis, intended to reduce pressure on Accident and Emergency departments.” These services can be provided by NHS trusts, third sector organisations or local authorities, or as a collaboration between these organisations. Typically, they operate outside of office-hours (5pm – 9am), and most allow service users to walk in without an appointment [8]. They are usually designed to be non-clinical services, providing a welcoming safe space for people in mental health crisis. Often, they are led by staff who do not have professional mental health qualifications [5] and may include peer-support workers and volunteers in their workforce [2, 18]. Crisis cafés are intended to be open to anyone who feels they need support, regardless of diagnosis. Typically, they are designed to be accessible at an earlier stage of crisis, before an individual is so unwell that they require inpatient hospital treatment [5]. They also aim to divert people from attending Emergency Departments when they are distressed but not necessarily in need of an intervention of a clinical type. This may prevent further escalation of crisis and addresses a gap in provision, where individuals are not yet severely unwell enough to qualify for crisis treatment, but may become so without prompt intervention [19].

The development of crisis cafés to divert people away from Emergency Departments has become an increasingly popular use for crisis care funding in England [8]. Between 2016 and 2019, crisis cafés were the most rapidly increasing service of all emerging crisis care models [8], and they were available in around one third (29%) of adult crisis resolution team catchment areas in England in 2019 [8, 20]. Grey literature has documented the development of some of these services [18, 21, 22]. One recent study suggests that having a crisis café as part of a local mental health crisis care system may be associated with lower hospital admission rates [23]. However, there is minimal published literature about their role in practice, how they function within the broader crisis care system, and what service characteristics and contextual factors affect how they function [5, 8], nor are we aware of publications from other countries describing similar models.

The current study was designed to address this research gap, investigating which factors affect whether crisis cafés implement their intended model and the barriers and facilitators to accessing crisis support through them. We sought to interview service managers from a range of crisis cafés across England, to seek their key perspective on their service’s development, and its current role and function within the local crisis care system.

This study built on interviews conducted for a previous project. The NIHR Mental Health Policy Research Unit (MHPRU) had conducted interviews with the managers of a range of innovative mental health crisis care projects, funded by the Department of Health and Social Care in England through the “Beyond Places of Safety” (BPOS) capital funding scheme, which provided infrastructure for development of services for urgent and emergency mental health care [24]. These included six interviews with managers of newly-established crisis cafes, previously analysed along with twelve other interviews to address questions regarding the crisis care system in general [25]. For the current project, we conducted six further interviews with managers of established crisis cafes, which had been running for longer than the BPOS-funded services. We analysed all 12 interviews afresh for this project, to explore the following research questions:

1. How are current services across England implementing the crisis café model, including their aims and subsequent operation?
2. What factors affect the implementation and access of crisis café support?

## Methods

### Study Design

We used a qualitative interview design to investigate the perceived aims of the model, how it functions in practice, and the barriers and facilitators to implementing the model as intended. The original BPOS study and our extension to the study with the additional crisis café interviews met Health Research Authority criteria for a service evaluation [26]. This was confirmed by the Director of the North Central London Research Consortium (NOCLOR) following review of the study protocols and interview guides.

### Research Team

BPOS interviews were conducted by UF, a researcher in the MHPRU. Crisis café extension interviews were conducted by HS, a post-graduate student researcher within the Division of Psychiatry (DoP). Alongside HS, a second coder GC, also a post-graduate student researcher within the DoP, PN, a PhD candidate and lived-experience researcher, and LM, a senior trainee psychiatrist and PhD candidate within the MHPRU, gave input into the generation of codes and their organisation into themes. Data collection and analysis was overseen by LM, SJ, a senior researcher and psychiatrist, BLE, a senior researcher and social worker, who are experienced in health service research, especially on crisis care.

### Study Sample

The study sample consisted of 12 service managers of crisis cafés from across England. We aimed to ensure our sample represented the wide range of crisis café services across the country.

For the initial BPOS interviews, the managers of all BPOS-funded crisis cafés were invited to participate. A purposive sampling method was used to recruit service managers for the extension interviews. As all BPOS interviews focussed on newer crisis cafés due to the timing of the BPOS funding, in this follow-on phase we aimed to interview service managers of more well-established services in the extension. To capture variation across the country, we approached service managers of urban and rural services, and services run by NHS, local authority and third sector organisations.

### Measures

A semi-structured interview schedule was developed by a working group of people with acute care, academic, practice and lived experience expertise. Topics adhered to implementation science frameworks, covering implementation of the intended model, perceived impact, barriers, and facilitators to implementation, [27, 28]. An adapted version that focused only on crisis cafés was developed with input from a lived experience expert. The final schedule (Appendix 1) covered: 1) description of the service, 2) service development rationale, 3) service implementation, 4) barriers and facilitators to implementation and access, 5) service effectiveness, 6) advice to new initiatives in development.

### Procedures

#### Recruitment

Participants were recruited either through details provided by the Department of Health and Social Care, or by email using contact details available on their websites.

All participants were sent a written information sheet to review, and their informed consent to participate was confirmed prior to being interviewed. They were informed that they would not be identified by name, role or service in the study findings.

### Data Collection

Interviews were conducted by video call, allowing people around the country to be efficiently reached. They were conducted by UF and HS between December 2021 and July 2022.

### Data management

All interviews were digitally recorded on Microsoft Teams, professionally transcribed, then checked for accuracy and removal of identifying details by the interviewers. Audio recordings were destroyed after analysis.

### Data Analysis

Data were analysed using a thematic approach [29] facilitated by NVivo12 qualitative analysis software. The analysis took an inductive approach and followed the 6-step model outlined by Braun & Clarke [29]. During the familiarisation process, authors read and re-read each transcript closely. HS generated initial codes by coding all interviews, with GC, PN and LM coding a proportion of the data and contributing codes. Following this, the coding frame was developed collaboratively and organised into themes. Initially this was a process of open coding, where there were no pre-defined codes, rather they were developed and modified throughout the coding process. The organisation of codes and generation of themes was performed by all members of the research team and was guided by the pre-specified research questions.

### Positionality

Researcher HS previously worked in inpatient mental healthcare, with clients in various levels of crisis. Preconceptions formed by treating patients who had recounted experiences of accessing crisis care may have shaped views in the current study. To include multiple perspectives on the data, a collaborative approach to coding was taken, with independent readings and interpretation comparisons of sample transcripts by GC, PN, LM, BLE, and SJ. BLE and SJ have led a programme of studies on crisis care and have given advice to policy makers in this area. SJ and LM also have clinical experience of working in NHS crisis services. PN has collaborated with NHS trusts, academic institutions and voluntary organisations developing and delivering community and in-patient based projects around mental health. A reflexive stance was employed by the research team, acknowledging our own preconceptions as academics and clinicians [30]. This included HS keeping a reflexive diary during data collection and using memos in NVivo during the coding process.

## Results

### Service Characteristics

Interviews were conducted with senior staff (service managers, CEOs, operations directors) from 12 services across England. Of these, eight were run by third sector organisations, two were run by NHS trusts and two were run jointly between the NHS and a third sector organisation. These services were spread across England: 5 were located in the North of England, 3 were located in the East or West Midlands and 4 were located in the South of England. There was a mixture of services located in urban areas (large towns and cities) (n=6), and services that served a more rural population (smaller towns and their surrounding catchment area) (n=6). The oldest service was established in 2014, and the newest services were developed during 2020, with a pilot phase in 2021/22.

### Qualitative findings

We aimed to explore the role of crisis cafes and what helps and hinders implementation of the intended model . Twelve themes were developed using thematic analysis, capturing service managers’ accounts of the intended role of the crisis cafes, the extent to which services were operating as intended, and the factors that were facilitated or hindered this. These themes are organised into three broad domains: i) The core role of the crisis café, ii) Factors that influence the effectiveness of a crisis café, iii) Key tensions for crisis cafés to consider. For each theme, illustrative quotes are provided A more detailed list of illustrative quotes can be found in Appendix 2. To ensure anonymity of individual services, illustrative quotes are not identified with location or provider organisation descriptions.

### The core role of the crisis café

#### Providing an alternative to Emergency Departments and hospitalisation

The main role of the crisis café, and the reason it was established within each care pathway was frequently cited as being an alternative to the Emergency Department (generally known as A&E in England), providing a place for people in crisis to receive mental health support, when medical intervention is not needed.

> *“The first outcome is trying to prevent people from going to A and E unnecessarily. The second one is we can better meet their needs outside, without needing psychiatric inpatient admissions, then it’s a success, and they’re supported safely in the community.” – Participant 7*

This alternative was suitable for people in acute crisis who required support and de-escalation, but not medical attention. Additionally, the crisis café was also seen as serving people in *‘lower level’* crisis, who use the service to prevent escalation to acute crisis.

### Improving access to crisis care

Whilst there was variation across services, most held core opening hours between 6pm and 11pm, and were open 365 days of the year. This was specifically so that people could access mental health support out of normal working hours when other avenues of support are closed.

Some services expanded their opening hours later into the night, or earlier into the daytime. This was reported to be motivated by a desire to be flexible for the people using the services, many of whom expressed a preference for extended opening hours.

> *“We have been doing a little bit of consultation with… our service users. They’ve been saying that they would use it in the daytime, actually, that it is something that they would like to make use of.” – Participant 10*

Other services, however, wished to remain operating purely out of hours despite requests to extend their services into the daytime. This was to encourage service users to make use of already existing support and develop their lives outside of mental health services during the day.

> *“Our feedback from those who use the service is excellent. They’re starting to say they’d like it in the day as well. [we say] “No, that can’t happen, because that’s a day hospital, and really, we want you in the day to be doing stuff.”” – Participant 4*

#### Providing someone to talk to, in a safe and comfortable space

A central aim identified for Crisis café was to provide service users with someone to talk to about how they were feeling. Even when a member of staff wasn’t available, being around other people and talking to other service users could help relieve people’s distress.

Creating an environment in which the service user feels safe, comfortable, and welcomed was a priority for service managers.

> A quote from a service user, relayed by the service manager: *“I was welcomed in and offered a cuppa and a safe space to sit and talk, a safe space being the thing I needed. I felt like I had walked into a chilled, normal, café environment with staff dressed not in uniform which really helped calm my anxiety.” – Participant 2*

The physical environment of the crisis café was seen as important in creating this welcoming atmosphere. Comfortable furniture, blankets and cushions contributed to the creation this sense of safety for service users.

> *“If no one has taken the time to give you somewhere warm and comfortable, how do you feel welcome? How do you feel listened to? How do you feel valued?” – Participant 4*

### Triaging effectively

Most managers described using a brief assessment to ascertain the risk level of service users. Sometimes this was structured by using tools developed for clinical services, at other times it took the form of a more informal conversation.

Risk assessment was seen as important, as crisis cafes don’t have the capacity to treat people who require immediate medical attention. This includes both physical healthcare and acute psychiatric attention. Managers described using a triage system to determine whether an individual fell within their remit of care. If the service user was deemed to fall outside of this remit, many tried to signpost to a more appropriate service, and sometimes helped them to access this.

> *“If there does need to be an escalation up to the crisis team, or to the liaison team at the local general hospital, which is where our psychiatric liaison team are based, we can arrange safe transport there for them.” – Participant 9*

### Improving crisis planning and people’s coping skills

Central aims identified for services were to provide a non-judgemental listening service and individualised practical and emotional support. A commonly stated aim was to improve people’s crisis planning and coping skills, enabling them to feel more able to manage their crises in future, or when the crisis café isn’t open.

Crisis planning included helping people identify where to get help in future (including from the crisis café) and to develop strategies to keep themselves safe.

> *“They’re telling us what’s causing them distress. We’re not giving advice, but what we’re doing is we’re listening, acknowledging, and sharing safety tips, strategies, techniques that somebody may want to try and to use, should they become in crisis in the future.” – Participant 11*

### Factors that influence the effectiveness of a crisis café

#### Accessibility

The importance of good accessibility was one of the main themes that we identified regarding factors influencing the success of crisis cafes in carrying out their intended roles. Challenges identified to accessing the crisis café included lack of available transport, safety issues, and cultural barriers.

Transport: Especially in rural areas, some individuals had no way to get home from the crisis café at night except calling a taxi, which many could not afford.

Some services had initially planned to provide taxis home for people, but many found this to be unfeasible in the long term with their budgets. Others kept offering free transport home using taxi services, but it’s “*it’s not something that we really shout about, because we don’t want it being abused*” – Participant 9.

Safety: Staff and service users could feel unsafe leaving crisis cafes late at night, and one manager reported that a service user had been assaulted on their way home. Some services had installed flood lights and CCTV outside their buildings. Others ensured they asked service users if they had a plan of how they were going to get home before they visited the café, encouraging them to be cautious.

Remote adaptations made during the COVID-19 pandemic had helped to overcome barriers to access for some who could not easily go out for a variety of reasons by enabling them to access support from home.

> *“There was a single parent that needed support, and one of the comments that she made was that, when we were able to give telephone support, she said, “I’m so pleased that you can do that, because I couldn’t access you before.” - Participant 11*

A need to do further work on accessibility was often identified. Some service managers acknowledged that there were communities within their local area that were still under-represented among their service users. Managers suggested that cultural barriers related to stigma and awareness around mental health services may be preventing some individuals from presenting to crisis cafés. They acknowledged that targeting identified communities and taking a more hands-on approach to overcome stigma within these communities may help to increase access.

> *“I do think we need to go out and be very much more hands-on in those communities to actually overcome the stigmas that a lot of people experience, and to make them aware of the service and that they can come to it. I think that’s a barrier, certainly for that particular group.” (referring to BAME communities currently identified as under-represented in their service)– Participant 8*

### Person-centred care

Co-production and consultation with service users emerged in most interviews as important to developing and delivering personalised care. Services had established service user networks and clinical advisory groups who had input into the initial design of the service, and later into the implementation and ongoing modifications.

Overall, this involvement was felt to have resulted in services delivering person-centred care designed to meet the needs flexibly of the local community, rather than to meet a set of stringent targets.

> *“We don’t have specific targets for having produced a certain amount of action plans, or safety plans, or that there are specific things that have to be done within certain timeframes, or any of those sorts of things. It’s very much, “Come in and use us in a way that is helpful.” – Participant 10*

Gatekeeping of services was kept to a minimum. A range of referral routes were used, including self-referrals, referrals from primary and secondary care, and referrals from carers. Most also operated an open-door policy, meaning anyone could walk into the service during opening hours and receive support. There were a range of service user preferences regarding referral routes, so allowing a service user to choose how and when they accessed the service kept the emphasis on person-centred care.

> *“Some people told us – they would only come if they could just walk in on the spur of the moment, and some people said they would never come unless they could phone up, and talk to people, and check they were okay. Other people wanted to be referred by their doctor or whatever, so we just felt that it was important that we provide that for people. So, that’s what we do.” – Participant 8*

Managers emphasised the importance of mental health crisis being self-defined. They aimed to assist anyone who felt they needed support due to their current mental state rather than to adopt a narrow definition of mental health crisis.

*“And there is a fundamental need to really shift the balance and understand that crisis, yes, is self-defined… And that it happens anytime, and my crisis is different from a neighbour’s crisis. So, we have to take people’s situations on a personal level.” – Participant 7*

Participants emphasised the importance of having very minimal eligibility criteria for their services. There were only three key exclusion criteria across most services:

1. Adults only. However, three services had flexibility to see 16-17 year-olds, they wouldn’t be immediately turned away.
2. People who needed immediate medical attention. This included people needing urgent physical healthcare (e.g. if they had harmed themselves or taken an overdose) or needing acute mental healthcare (e.g. someone who appeared to require immediate inpatient admission). These individuals would instead be referred to A&E, and crisis cafés would assist them in getting to the hospital by liaising with emergency services or calling a taxi.
3. People heavily under the influence of drugs or alcohol or exhibiting aggressive or violent behaviours. This exclusion was based on their current state, with intoxicated service users told that they would be welcomed back once sober and able to engage with support.

Crisis cafés were generally intended to embody the ‘No Wrong Door’ policy, recognising that mental health is complex and may need input from multiple services within the community. Thus services had formed links with a variety of local services, including for example substance misuse services.

### Relationships with other services

One of the most prominent themes we identified was the importance of the relationships that crisis cafés had with other such services.

Relationships had begun at the consultation phase when the services were still being designed. Service managers recounted visiting other crisis cafés to understand the models that they used, as well as advising new initiatives on learnings from their own services. One service manager took members of the local Clinical Advisory Group, comprised of service users, to visit other services and get their views on how they operated, which then influenced how they designed their own service.

> *“So what we wanted was, we wanted the hybrid model basically. So we liked bits of [service 1], and we wanted bits of [service 2], so that’s what we developed.” – Participant 3*

Being integrated within the local mental healthcare system and with other community services also influenced how the crisis cafés operated. Having working relationships with other care providers meant that they could take referrals for people who did not meet criteria for other services and offer them alternative support, helping to ensure that service users don’t slip between the cracks. Some services could refer directly from the crisis café to further resources in the community with the understanding that the service user would be seen by them.

> *“So, we’re trying to connect the idea behind working together with third-sector organisations, is to try and connect with community-based assets, to try and link in people with things that are local to them, and support.” – Participant 7*

When services were less integrated in the existing pathways, it presented problems. In a newer service significantly impacted by COVID-19, the managers described a more stand-alone service that struggled to get referrals from outside their service. Operating out of hours was sometimes a barrier to establishing relationships with daytime services: managers recommended initiating these working partnerships in the design phase, and reported that persistence was key to creating effective partnerships. .

> *“That’s what we’re talking about, the crisis care system that we were meant to link in, as we said, but it didn’t happen. So we obviously linked in with them when we had to refer, but that’s what we were doing anyway. So, yeah, is hinder the right word? It just didn’t happen.” – Participant 6*

### Staffing

A further facilitator to success in achieving intended goals was sufficient staff with the right skills for the job. Crisis cafés employed a wide range of staff. Most did not have formal mental health qualifications and came from a range of backgrounds. Job roles included crisis support workers and peer support workers; some had counselling backgrounds, whilst others came from health and social care and other areas of pastoral support.

Service managers emphasised the importance of adequate staffing for a successful crisis café. Making sure there are enough staff on shift to provide support for the level of demand was important and having a bank of staff you could call upon when running short of staff helped meet this need.

> *“However many staff you think you need, add at least two more” – Participant 11*

Managers reported that service users valued having staff with lived experience of mental health crises within the crisis café. Often, they wanted someone to be able to listen and empathise with their situation, not so much to give them solutions, but rather to validate their feelings and support them during a distressing time.

> *“People are often surprised because it’s not someone with a string of qualifications who’s sitting there, going, “I know best because I’ve studied this for 17 years.” It’s, “I live like this every day, like you live, and this is how I find it.”” – Participant 8*

Several managers mentioned the importance of having senior colleagues with lived experience who influenced policy and practice of the crisis café. Two services were survivor-led, with leaders’ experiences of the mental health system influencing key decisions made throughout the organisation.

Service managers saw bespoke training for working in the crisis café as essential, giving staff the confidence and ability to handle difficult situations and provide the best support to service users. Types of training included Mental Health First Aid [31], Prevention and Management of Violence and Aggression (PMVA) [32], and Intentional Peer-Support [33].

> *“Make sure that actually staff are receiving the training and support to deal with the fact that, although it’s a non-clinical service, they are at times dealing with some very difficult circumstances.” – Participant 10*

Even with training, staff inevitably encountered situations that they found worrying or distressing. Making sure staff felt safe and able to do their job was seen as important, including physical safety. One café had installed floodlights in front of their building to ensure that both staff and service users felt safer when leaving the service late at night.

Some services could access clinical supervision with colleagues from crisis teams which provided reassurance and guidance for more complex cases, and aided staff development. Managers reported that good supervision was especially important when employing lived-experience professionals. Staff who draw on their own experiences of mental health crises may be at risk of becoming distressed themselves. Specific provisions needed to be made so that staff have time for debriefs and supervisions within their working week, to ensure a dedicated space where they can process what they have experienced.

> *“Supervision, the reflective practice, the debriefs is really important, […] so people can think through and process all the thoughts and feelings that they’ve experienced in the work that they do.” – Participant 12*

### Key dilemmas for crisis cafés to consider

#### Open door policy versus managing demand and the practicalities of referrals

Many services took pride in accepting walk-ins as part of their service provision, seeing this as key to accessibility. However, this open-door policy could become an obstacle. when it came to managing the demand for the service.

> *“Until 11:30pm you can walk in, unannounced, and just say, “Can someone listen to me?”” – Participant 4*

Staffing levels, physical space and more recently COVID-19 restrictions also limited the support that services could offer. With increasing demands for their support, some services with an open-door policy had found themselves having to turn people away.

> *“It is getting now where we do have people ring up and we can’t necessarily always see them that day. Which is always quite disappointing for people, when they’ve had the courage to pick the phone up.” – Participant 5*

One service found that a more restrictive self-referral procedure was needed to manage this demand, where service users had to ring up and be offered an appointment for that night.

> *“We created these personal time slots, so that’s where the two hours came from, and then the one-hour telephone support as well.” – Participant 11*

#### Risk assessment and the remit of care in the context of offering a non-clinical service

A further dilemma apparent in many accounts of services was between sufficient risk assessment and offering an essentially non-clinical service. Being distinct from clinical services was often seen as positive, offering greater flexibility and a more human and direct response.

However, considerations of risk and liability could not be entirely put aside and were seen as relevant to the welfare of service users:

> *“No, we don’t have a governance duty of care to them. But we’re not mean. So, we will try and make sure, without claiming responsibility for people.” – Participant 4*

Stated principles for some services were that they had a duty of care only while service users were inside their facility. For others, the remit of care extended beyond the opening hours of the service, for example checking in with service users during the daytime or making follow-up calls to other services they referred to.

> *“We do have a number of hours during the day where our team manager, she will make follow-up calls to people as well. So, we will check in with people and our daytime service would welcome those people in as well.” – Participant 9*

Introducing stricter risk management procedures risked making non-clinical services feel more akin to their clinical counterparts, potentially a move away from their current service ethos and goals.

Interviewees noted that currently service users aren’t afraid to talk to crisis café staff, because they believe compulsory detention under the Mental Health Act would not be initiated at the service. .

> *“Sometimes people need to talk about suicide and how they feel about it, without it becoming a, “Quick, let’s 136 this person,” before they can say any more.” – Participant 8*

> *(quote refers to the use of a “section 136”, a power held by the police in the UK to detain a person under the Mental Health Act for up to 24 hours to facilitate transfer of the person to a place of safety, such as an Emergency Department, and holding there for assessment)*

#### Visibility and increasing awareness of the crisis café versus avoiding perceived stigma in the community

A further identified dilemma was between visibility in communities and privacy/avoidance of stigma. Crisis cafés attracted more users by word spreading throughout the community and managers valued being embedded within the community. However, they were also aware that stigma may act as a barrier to accessing care: people might not wish to be seen going into a crisis café if other people knew that meant they were accessing mental health support.

Some crisis cafés had tried to resolve this potential conflict between visibility and privacy by making their premises geographically accessible, with access to public transport links nearby but without making it obvious that they were mental health services.

> *“They wanted it to be accessible, but they didn’t want it to be on a high street or something, because they didn’t want someone they knew to see them going in.” – Participant 8*

## Discussion

This study investigated the crisis café model of support in England, through interviews with service managers from crisis cafés across the country. Our analysis identified five core aims of crisis cafés that form the basis of how they operate. Four main factors were seen as influencing the success of services in meeting these aims, relating to accessibility, person-centred care, integration into local services systems and quantity and quality of staffing. Finally, several dilemmas emerged, where services had to consider important trade-offs between priorities for their services. .

### Findings in context

There are parallels between the four main factors identified as associated with success, and key ingredients of high-quality crisis care identified through research. In a fidelity measure developed for crisis resolution teams (CRTs), important components of the CRT model fell into 4 clusters: referrals and access; content and delivery of care; staffing and team procedures; and timing and location of care. Aspirations for crisis cafes that were congruent with these criteria included rapid response, accessibility, individualising care and delivering services away from institutional settings [34]. Across voluntary sector crisis care provisions, accessible location, good availability, flexible eligibility criteria and good connections with the rest of the crisis care system referral routes were all identified in a national investigation in the UK as playing an important role in improving access to crisis care tailored to individuals [2].

Across crisis services, a point of contention is the definition of mental health crisis used. In this study, multiple services emphasised the importance of crisis as a self-defined concept, which was reflected in how the services were designed. One person’s crisis may look different to another’s, but the support that crisis cafés offer should be able to cater for every person’s definition of crisis (within the scope of their eligibility criteria). Several key papers on crisis care provision have examined the definitions of crisis [1, 2, 35]. Pragmatic service-oriented definitions, self-definitions, risk-focussed definitions and theoretical definitions have all been explored [35]. Commonly in voluntary sector led crisis services, the importance of self-definition of crisis is emphasised [2], alongside its role in validating service users’ experiences and promoting acceptance. This links back to views expressed in our study, where service managers wanted to shift the balance towards understanding crisis on a personal level and be able to offer support to anyone who felt they needed it, without requiring external confirmation that the individual meets certain crisis criteria. This contrasts with the more restrictive operationalisation of mental health crises in crisis resolution teams, which aim to focus on crises sufficiently severe to be close to the threshold for inpatient admission: this difference in scope may allow crisis cafes to intervene helpfully before crises become very severe, but the flexibility of their definition of crises also makes managing demand challenging.

Within all themes, the idea of flexibility within services was emphasised. The NHS long-term plan recognises the importance of tailoring provisions to meet local needs, including identified ethnic and cultural health inequalities and the availability of services within the local pathway [36]. Across interviews there were a range of different types of crisis cafés and implementation strategies. Discussions of key dilemmas suggested that there is not one gold standard way to operate a crisis café, but rather different trade-offs to be made. Decisions about how to find a balance between competing priorities are informed by the identified needs within the community, consultations with service-users and lived-experience experts within the area, and the structure and operation of the existing local crisis care pathway. The idea that fidelity to a model might include flexibility and tailoring to local context has informed implementation for strategies for other mental health care models, and may be applicable to attempts to further define the crisis cafe model for future implementation and research [37]. This concept could be explored for crisis cafés in the future.

### Strengths and limitations

This is the first study to explore in-depth crisis café model of care across a range of services. The interviews conducted with senior staff within these services provide good insight into current practice, and the themes that emerged from our analysis have potential to inform further service development and research. Our recruitment strategy may have limited the perspectives we heard from during the study. By recruiting current service managers, we were unlikely to hear from crisis café initiatives that hadn’t succeeded. From previous case studies [2], we know that some crisis cafés have closed after their initial pilot phase, for reasons including under-use. Insights into these services could have provided an alternative view into what factors influence the running of a successful crisis café.

Similarly, our sample may have over-represented positive views of crisis cafés and their role within the wider system, as all interviewees led such services and had an interest in securing further funding. Further research in this area needs to include the perspectives of a wider range of stakeholders, and co-produced research into service users’ perspective on these services remains a critical evidence gap.

Cultural barriers to accessing crisis café support were brought up in several interviews, in common with other studies in the crisis care literature [2, 35, 38]. Interviews with managers provided only limited opportunity to explore this. Future research should include the perspectives of communities identified as currently under-represented in services.

Finally, all the included services were in England, so we have no basis for commenting on the model’s applicability elsewhere. However, crisis cafés exist in other countries, like Australia [39], with some even modelled on examples from England [21, 40]. Differences in local and national crisis care system may mean that the way services operate and challenges in implementing the model vary between countries.

### Future directions

Research with service users themselves is needed to gain a further understanding into what makes an effective crisis café from the perspective of people who use the service: co-produced or service user-led studies are preferable to ensure that the questions that are important to service users are addressed.

Some individual crisis cafés had started evaluation projects to quantify their impact within the local mental healthcare pathway, for example investigating changes in numbers of Emergency Department presentations, or numbers of people for whom they believed crises had been cases successfully de-escalated and admissions avoided [22, 41]. However, further quantitative investigation is needed across multiple sites with appropriate comparators to determine whether crisis cafés are in fact achieving their aim of providing an alternative to A&E when in crisis, and to understand which groups of people in which contexts are best served by them. Comparisons of outcomes for people using crisis cafés with those of other more established forms of crisis care, like CRTs are of interest [5, 6, 35], as well as investigation of the characteristics of people using these services and identification of communities who are not currently engaged by them.

### Conclusions

Crisis cafés are currently growing in popularity and are also significantly under-researched. This qualitative study is the first to investigate the crisis café model of support in depth. We identified a core set of aims for crisis cafés that inform how they operate in practice. Several key factors influence the effectiveness of service provision, by facilitating effective care, or acting as barriers against it.

These factors are similar to key ingredients identified in other forms of crisis care. Furthermore, there are key trade-offs that services need to consider when designing and running their service. Further research with other key stakeholders is needed, as well as quantitative evaluation of service operation and outcomes.

## Supporting information

Appendix 1 - Interview Schedule

Appendix 2 - Supplemental Table of Illustrative Quotes

## Data Availability

All data produced in the present study are available upon reasonable request to the authors

