## Appendix 1 - Interview Schedule for "A qualitative investigation of crisis cafés in England: their role, implementation, and accessibility"

**Appendix 1: Crisis Café Extension Interview Schedule**

### MHPRU – BPOS Project Managers' Interviews

### Adapted Interview Schedule – Crisis Café Extension –25/03/2022

**Project information**

**Name of crisis café:**

**Name of provider organisation (e.g. NHS Trust, voluntary sector organisation):**

**Respondent's role/job title in the crisis café service:**

**When was it established?**

**1. Please can you briefly describe the crisis café that you work in.**

Additional Questions/Prompts:

- *What are the aims of your crisis café?*
    - *Additional prompt if this seems vague: what do you offer for people who attend the crisis café? What are the aims of this?*
- *What is the target population of your crisis café?*
    - *Are there any exclusion criteria for potential crisis café users?*
- *How do people access your service? (e.g. open door, call ahead, referral/self-referral?)*
- *What types of staff make up your crisis café? What roles do they play in your care delivery? ***

Additional Questions for well-established crisis cafés:

- *Has the way your crisis café operates changed over time?*
- *Are there any changes to your service that you have needed to make? What were the reasons for this?*
- *Have the aims of your crisis café changed over time? How so?*

**2. What was the rationale for the development of your crisis cafe?**

Prompts:

- Were service users/people with lived experience involved in the development of your crisis café?

**3. Has the crisis café been implemented successfully?**

Prompts:

- *Take up: how many people used the service,* ***were they the target population/clinical group****?* [Adoption and penetration]
- *What is the current demand for your service? How is the crisis café equipped to manage this?*
- *Has the service been set up and delivered as intended* [feasibility and fidelity]
- *How has the service been received by staff (within the service and in other crisis care services) and service users?* [Acceptability and appropriateness]
- *Has it been delivered on budget? Do you think it is good value for money?* [cost]

- *Will it continue as planned longer term?* [sustainability]

**4. What has helped and hindered implementing the crisis café?**

Prompts:

- *The nature of the crisis café itself* (e.g. clarity, simplicity?) [Intervention]
    - *What activities/support is offered? Does the café cater to people in acute crisis, or lower risk individuals?*
    - *Transport (especially in cafes serving rural areas) and language barriers?*
    - *Opening hours – any feedback from service users?*
- *Attitudes, skills and behaviour of staff running the service* [Staff]
    - *Diversity of the workforce, are there peer support workers with similar lived experiences, or a range? Do they employ staff from a range of cultures? Is English the only language spoken by the staff?*
- *The local crisis care system (fit with other crisis services, attitudes and level of support from managers* [Inner setting]
    - *For crisis cafés run by voluntary sector organisations, do they work in partnership with an NHS trust, or are they stand-alone? Does this affect what support users can access?*
- *The impact of the Covid-19 pandemic on the implementation of the crisis café* [Outer setting]
- *The wider context (needs of the local population/service users, any impact of local or national policies)* [Outer setting]
- *The process for starting up and embedding the crisis café (extent and success of planning, engaging stakeholders, executing plans)* [Process]
- *Extent and impact of any coproduction with service users in designing and implementing the project [Process]*

**5.How effective has the crisis café been in improving patient care and outcomes**

Prompts:

- Has any evaluation been conducted?
- Has any equalities impact evaluation been conducted?
- Is there any available routine data which could help assess the impact of the crisis café? – e.g. number of presentations to A&E vs. crisis café
- Perceived impact of the crisis café?
- Any unintended consequences from the programme?

**6. What advice would you give to other NHS Trusts or health care providers who were thinking about a service initiative like yours?**

**7. Are there any available reports or summary data, which you would be happy to share and which include no personal data, regarding changes in service use and patient flows or service outcomes within the crisis café or the local crisis care system, which could help us understand the role and impact of the crisis café?**
