## Appendix 2 - Supplemental Table of Illustrative Quotes for "A qualitative investigation of crisis cafés in England: their role, implementation, and accessibility"

### Appendix 2 – Additional Illustrative Quotes

| Core Role of the Crisis Café | |
|---|---|
| Effective triage – assessment and referrals | "And we could then refer on to the CMHT or a home treatment team to be able to offer follow up support, or for them to come back to The Retreat the next night to have kind of further ongoing support." – Participant 3<br><br>"If there does need to be an escalation up to the crisis team, or to the liaison team at the local general hospital, which is where our psychiatric liaison team are based, we can arrange safe transport there for them." – Participant 9 |
| Improving crisis planning and people's coping skills | "We try and send everybody off with a plan. We try and we use it, so it's quite… If somebody comes in and they're in, you know, they may come in in a very difficult position. They may feel they're not sure whether they can keep themselves safe. They may say they can't keep themselves safe. Then staff will work with them over the course of an evening, and that may change their mood." – Participant 8<br><br>"Then, I think, in terms of the sort of support that we provide, it is very much that listening ear and signposting. We do safety planning with people, as well, if that's required when they come in." – Participant 10<br><br>"What we're doing there is we're being guided by the individual. They're talking us… They're telling us what's causing them distress. We're not giving advice, but what we're doing is we're listening, acknowledging, and sharing safety tips, strategies, techniques that somebody may want to try and to use, should they become in crisis in the future." – Participant 11 |
| Operating outside of normal working hours | "So, originally, it was just open for evening sessions a week, and now- Well, hopefully, in the coming weeks. It has already extended its opening to seven days a week, but we're going to be further extending the hours. So, it should be open from midday until midnight." – Participant 1<br><br>"They were struggling to- the times when they were struggling were mostly out of hours, the weekends, and Bank Holiday weekends, like we're coming up to a long Bank Holiday weekend, it becomes quite difficult, even appointments in the week, to see psychiatrists, to see their CPNs would be quite difficult." – Participant 7<br><br>"We have been doing a little bit of consultation with… our [area] service users. They've been saying that they would use it in the daytime, actually, that it is something that they would like to make use of." – Participant 10 |
| Providing someone to talk to, in a safe and comfortable space | "We're going to have some service user involvement around, again, physical appearance of the inside of the building, just to make sure that we're not turning it into some God-awful NHS spec clinical space. We still want it to feel- We want it to be safe, but we want it to feel very, very welcoming." – Participant 1<br><br>"So, it's like we're not like a clinic, or anything. The way that the communal area is set out, part of it is set out as a café. So, we've got round bistro tables and chairs. The other part of the room is set out with tub chairs and coffee tables, and things. So, you're not walking into something that looks clinical. You're walking into somewhere that is welcoming." – Participant 9 |

| | |
|---|---|
| | "The crisis cafes do is provide – not for everyone but provide for a lot of people – what they are looking for, which is really a safe space to go that they feel comfortable in, safe in and they feel that their distress is heard, acknowledged and understood without being assessed or asked a lot of intrusive questions about this, that or the other." – Participant 12 |
| Providing an alternative to A&E | "It's something else that we can use to divert people from our place of safety or emergency department, so that they can receive their crisis care in a sort of non-clinical, much more user-friendly third sector kind of environment." – Participant 1<br><br>"Where, you know, they don't have to be sitting and waiting for hours in A&E in order to be triaged. Because they can literally walk in the door there and be seen within ten minutes or so at the Crisis Café and have that one-to-one bespoke conversation." – Participant 2<br><br>"We're non-clinical. We don't have any kind of clinical staff members in our service and so, yes, we are there to try and provide an alternative, as well, to A&E. For people who don't require a medical intervention" – Participant 10 |
| Factors that influence the effectiveness of a crisis cafe | |
| Accessibility | |
| Cultural barriers | "It would be better if it was used by the older generation, as well. But for some reason, they don't seem to attend. I think it would be good as well to make sure that it's more diverse, and to have people from other ethnic backgrounds to attend, but they don't tend to use that service. So, for instance, [town] is in a military town, with a Nepalese community. And they don't generally tend to use this service as much. So, on that basis, maybe it should target better, in terms of diversity across age, and ethnic origin." – Participant 7<br><br>"I think the thing I would like to address, when things go forward, is around diversity *[...]* we're not getting a lot of people through the door from different ethnic groups *[...]* I think my feeling is we have to go out and work with them in their communities, not expect them to come to us, but at the moment, in the pilot phase, there isn't the capacity to do that, because we've been learning so much about just running the service and growing it." – Participant 8 |
| Transport | "Another barrier we've found is people actually accessing the service. So, we're looking at how we can increase kind of patient transport offer to the service as not everyone at this has access to cars. When you're in a mental health crisis you shouldn't really be driving anyway because it's not safe for yourself and other people and, you know, public transport isn't always an offer for everyone." – Participant 2<br><br>"Because the reality is, unless you're in a city centre, which we're not, people don't want to come out, because public transport gets worse and it's dark. And they just don't like that level of travel at night in more rural areas." – Participant 5<br><br>"We used to have a taxi service. Unfortunately, that just wasn't sustainable, given the numbers of people that we support. We would spend thousands and thousands on taxis" – Participant 11 |
| Safety Issues | "For example, the lighting being an obvious thing, and making sure that the front door was very secure. Because the front door of the building, as it was when we took it over, you could have pushed it open, really. And I think it's very important for people to feel safe and secure in their working environment, isn't it? And I think the lighting issue, in particular, given the location" – Participant 1 |

| | |
|---|---|
| | “I can, maybe, remind people that this is a night-time project and not everyone is lovely.” – Participant 4<br><br>“So, we can alert the CCTV operators that we’ve had somebody come to our crisis café that we can’t allow in. We can give them a description and then CCTV can pick them up and make sure that they’re alright.” – Participant 9 |
| Remote adaptations improved accessibility | “Public transport is not very good once you get out of those commuting hours, so actually, for some people, it has provided a really useful alternative so that they can still access that support. I think people have got used to receiving support in alternative ways, as a result of COVID. It forced people to have to be okay with it.” – Participant 10<br><br>“There was a single parent that needed support, and one of the comments that she made was that, when we were able to give telephone support, she said, “I'm so pleased that you can do that, because I couldn't access you before.” That was an eye-opener for us. Where we’d assumed that face to face was the best, actually there were a lot of people that we realised couldn't access our support, because couldn't leave children at home on their own. Maybe there was a disability there that meant that they couldn't leave their house and that kind of thing, fear of leaving the house, crippling anxiety.” – Participant 11 |
| Person-centred care | |
| Co-production and consultation of service users | “So, the whole of our safe haven project, if you like, from start to now, has been coproduced. So, we had service users involved in the very early days of developing the safe haven. We have run, since it has been open, a number of service user and carer events and focus groups. We take service user feedback for every attendance, if people are happy to give it. And we utilise that in a variety of different ways, including our ongoing service improvements and developments.” – Participant 1<br><br>“They [service users] were kind of utilised in order to help devise a name for the service, devise how the space should look. So, they provided input on colour schemes to use, imagery to use on the walls and things like that. So, yes, there was membership engagement.” – Participant 2<br><br>“Everyone who works for our organisation has had some form of experience of past trauma, not necessarily be involved in mental health services, but a number have. And on the board, we have to have – I think it's over 50% of board members who've had their own experience of crisis.” – Participant 12 |
| Designed to meet the needs of service users, not target driven | “Because we're person centred and person focused, then we’re not really worrying about targets and things. We're worrying about the person, so that's also a different approach and makes it very positive.” – Participant 8<br><br>“I do think some of our service users, as well, find it quite useful, the fact that it is very much led by them. We don't have specific targets for having produced a certain amount of action plans, or safety plans, or that there are specific things that have to be done within certain timeframes, or any of those sorts of things. It's very much, “Come in and use us in a way that is helpful.”” – Participant 10<br><br>“We're quite lucky in the sense that, formally, we don't have any kind of formal key performance indicators for the Sanctuary“ – Participant 11 |

| | |
|---|---|
| No gatekeeping - range of referral routes, encouraging walk-ins, crisis is self-defined | “You didn't need an appointment, so people could just turn up, relatives could just turn up, and they could sit and have a coffee or whatever while they waited for somebody to be seen. So there was no appointment system in place.” – Participant 3<br><br>“I remember when we started and they were like, “What is the referral process going to be? Is there going to be a referral process?” I went, “Well, that’s idiotic, genuinely. I have a walk-in crisis team. I can’t have my lower-level crisis support and you need a referral for it. That doesn’t make sense. It has to be more accessible, not harder.”” – Participant 4<br><br>“Some people told us – they would only come if they could just walk in on the spur of the moment, and some people said they would never come unless they could phone up, and talk to people, and check they were okay. Other people wanted to be referred by their doctor or whatever, so we just felt that it was important that we provide that for people. So, that's what we do.” – Participant 8<br><br>“It's very much about, if somebody feels as if they need that support, then we're there and we're able to offer it, there's no… Yes, we're a mental health Crisis Café, but the crisis is service user defined, so it's if they feel as if they need that support, as opposed to requiring any kind of other referral or external confirmation about actually whether or not they're in crisis.” – Participant 10 |
| No wrong door policy, minimal eligibility criteria | “In the Safe Haven, we don't turn away people, obviously. I think, earlier on you asked me about criteria. We don't turn anyone away. The only reservations we've got is around people who are absolutely under the influence of alcohol who are violent towards members of staff, and furniture, and property, etc.” – Participant 7<br><br>“We’ve been very, very clear right from the get-go that, if somebody needs the medical attention… But having said that, we would probably ring them a taxi and get them up to A&E anyway. Again, not allowing people in that are intoxicated for whatever reason. Again, that’s health and safety, and the safety of the staff and other clients.” – Participant 9<br><br>“But the crisis cafes could still support anybody. So if someone arrived on their door in an acute state of mental distress, they could still support them, but as it's a social space, the emphasis is more on a social space of safety and warmth and security.” – Participant 12 |
| Relationships with other services | |
| Consultation with other services during service design phase | “We have been approached by other areas and we have offered advice and support. In the same way that, when we were thinking about it, actually, we visited a couple of other places that had safe havens, including [service], who have got a really, really excellent provision there.” – Participant 1<br><br>“Come and visit the service really, and talk to people who've been involved in it really, because I think some of the pitfalls, some of the learning from us surrounded how you can kind of challenge some of those belief systems that some people have.” – Participant 3<br><br>“I think one of the things that were good was doing the consultation first and making it quite a wide-ranging consultation, because I think people were then… A) It raises awareness of the fact there's going to be a Crisis Café, and b) also it gets people invested in it. I think, if you're going to have people with lived experience working in a service, you have to understand the challenges that that gives.” – Participant 8 |

| | |
|---|---|
| Embedded within the community | "Yes, [crisis café] as a whole, is really embedded in the community. Part of my role is I sit on the majority, if not all of the mental health committees and forums across [the county]. So, although we're quite a small charity, we don't work in silo, we're very much about partnership working." – Participant 9<br><br>"We keep up to date with things that are going on in terms of other community-based support services, because one of the things that we need to do is actually we work really hard to try and empower the people who come in to see us, to be in charge of their own recovery journey." – Participant 10<br><br>"Having those kinds of agreements around you is really helpful, so knowing who the local person is in the housing department, because the risk of homelessness is quite a big factor that we see in the Sanctuary, so knowing that you can contact someone and say, "This person came in to see us last night. They're struggling. Can you give them a call?" and it will be done." – Participant 11 |
| Integration within the existing mental healthcare pathway | "I think the [crisis café], obviously, works best when it's part and parcel and it's well integrated into the local mental health services, particularly the trust. Because it just makes more sense to really have a streamlined, and a good communication flow, particularly when people are presenting in communities. We just need to make sure that the pathways are clearly defined, and well understood." – Participant 7<br><br>"Yes, we work really closely with our local crisis team. We hold locally what we call the 'focus network', which is a multi-agency group, and we meet monthly. Each agency can bring a client or clients to the network who we feel are presenting at multiple places and causing us all a lot of work. Then we can hold a meeting for that client with the client present." – Participant 9<br><br>"We keep some kind of notes and things. So, after someone has been to see us, we'll write that up. That information goes back to the First Response Service, which will then make its way on to their GP notes and things. *[…]* So, we're all working together, rather than Joe Bloggs having to tell his story again, and again, and again." – Participant 11 |
| Referrals to other services | "A number of options might come out of that assessment. One might be that the person requires secondary services, and what type. So, if they're already known to services, maybe we could signpost back to there, and if they're not acutely unwell, we may refer them to the same HRS. And if they are acutely unwell, and we think that they require acute care pathway intervention, we will liaise with the home treatment team, as well, to see whether we could do one or two things, which is to offer home-based treatment as an alternative in the first instance." – Participant 7<br><br>"Basically, the idea is that, I don't know, Joe Bloggs comes in to see us in the Sanctuary and he shares, I don't know, that his drinking is, maybe, spiralling out of control and that's worrying him a bit. We will ask Joe Bloggs: "That sounds like something you might want to tackle. Would you like us to make a warm handover to CGL?" Then they guarantee that then Joe Bloggs would be contacted the very next day, so it reduces that need or that risk of someone slipping through the net or saying, "Here's a telephone number for you to call," and then they never call that number. It's too scary or whatever." – Participant 11 |
| Staffing | |
| Employing staff with lived experience | "I think, because people have lived experience, our staff, which can have its own challenges, obviously, but because they have that, because they can sit and talk to someone who might be on the same medication as them or might have the same diagnosis as them and who has been through things, who has also got a child with autism or also has other things that they're experiencing, then it's really quite reassuring." – Participant 7 |

| | |
|---|---|
| | "A lot of our staff have their own direct experience of mental health challenges themselves. So, what they will do is they will utilise their own experiences of challenges, share tips and strategies that they've used in the past, what worked, what didn't. That helps evolve those support conversations around what people might like to try in the future." – Participant 11 |
| Staffing constraints | "We've limited that to how long they can have a word in private for, because of practical reasons, really. Not wishing to be rude to people, but when you are in that Zoom environment, you can't do that, because you've got another 12 people sat chatting amongst themselves." – Participant 4<br><br>"So, yes, there are not enough people and there are not enough staff to do everything we want to do." – Participant 4<br><br>"We also find that retention of staff can be difficult. People can come along and think that it will work quite well if they have a family, for example, but then the reality is that actually you're never home at teatime and bedtime, and if you have children and those sorts of things." – Participant 10 |
| Support available for staff | "Basically, if we're preaching that we're supporting our service users from a mental health point of view, surely we should be doing it with staff." – Participant 6<br><br>"Sometimes the things that people say are quite traumatic. Although we do support, we do co-supervision, we do supervision, I think from time to time it benefits people to be completely outside the organisation and just have a sound-off. I think that would be really useful to build into any service." – Participant 7<br><br>"We are building in additional hours that are what we call 'off-shift' hours. *[...]* what you then find is that actually you are in a constant struggle to find protected times for things like one-to-one supervision, and reflective practice support, and opportunities for networking and doing some of those other bits and pieces. *[...]* I think building in some of those additional hours within the staffing structure is really beneficial and would be really important to just enable some of that stuff that needs to happen." – Participant 10 |
| Training of staff | "Mental health first aiders, we've still got them all trained up and they're still delivering that sort of service really, to the people we support already, if that makes sense." – Participant 6<br><br>"It also allows us, I think, to manage situations as they arise, a lot more. We are mindful of risk. I've given the staff a lot of training in risk, and I've given them… We've had PMVA training and skills, particularly because they're all – well, most of them are – women. (Laughter) You just feel, '"Okay, they need to be able to handle these situations," and give them the confidence to handle those situations if they should arise" – Participant 8<br><br>"There are times where there are some very difficult situations that our staff are having to deal with. So, although it's non-clinical, it doesn't mean that actually the intensity of some of the work that you're dealing with isn't there. So, I think it would be about making sure that people were aware of the breadth of service users and the issues that we can then find that they bring that they will need to equip staff to be able to deal with. I don't know if there's anything else." - Participant 10 |

| Key tensions or considerations for crisis cafes | |
|---|---|
| Open door policy vs. the practicalities of referrals | “We are just saying things like, “Maybe ring before you come. Because if it’s busy, you won’t be able to come in. So, just check out that you can just walk in. I haven’t got anywhere for you to stand. It’s the centre of town, opposite quite a-”” – Participant 4<br><br>“What we found was that, yes, it was lovely for someone to be able to spend, let's call it, a whole shift with us, if you like. What we actually identified is there were people that were in crisis who weren't able to come in, because we were spending such a long time with one particular person, sort of thing.” – Participant 11<br><br>“We basically said, “Look, this is the challenge we've got. We’re supporting people but know we've got all these other people that we need to support as well, but can't. What can we do?” Anyway, the upshot of that was that we created these personal time slots, so that's where the two hours came from, and then the one-hour telephone support as well. That has worked really well. We've found, on average, that a face-to-face support session would probably last about 1 hour 50 minutes. That's generally how they are. A telephone call is normally about 50 minutes as well, so having the hour and 2 hours just gives us enough time. So, that was one of our major changes, and that has worked really well ever since.” – Participant 11 |
| Risk assessment and the remit of care vs. being a non-clinical service | ““Don’t you have a duty to her?” I went, “Hmm, yes and no. So, the answer is yes, while she’s in our facility, but she made her own way there and it’s December, and it was dark when she arrived.” – Participant 4<br><br>“I think in the NHS, I think we'd be much more procedurally bound and risk bound, whereas actually we can take decisions that we feel are right.” – Participant 8<br><br>“If I tell people that they need to be cautious on getting there, then I’m saying, “We know it’s dangerous for you to travel at night.” It’s just so ridiculous and so complicated.” – Participant 4 |
| Visibility of the service vs. avoiding perceived stigma from the local community | “Again, barriers, I think it’s that stigma, isn’t it? It shouldn’t be there, but it is. “Oh, am I going to be judged? Is it really for me? Can I go? Do I want other people to know that I’ve been there?”” – Participant 9 |
